# Physical Activity Moderates Inflammatory Gene Expression in Rheumatoid Arthritis

**DOI:** 10.1101/2021.08.27.21262729

**Authors:** Sarah L. Patterson, Shenghuan Sun, Dmitry Rychkov, Patricia Katz, Alexandra Tsitsiklis, Mary C. Nakamura, Paula Hayakawa Serpa, Charles R Langelier, Marina Sirota

**Author notes:** Equal contributions (Sarah L. Patterson, Shenghuan Sun, Charles R Langelier, Marina Sirota). **Corresponding Author:** Sarah Patterson, M.D., Division of Rheumatology, University of California, San Francisco, Box 0500, San Francisco, CA 94122. **Competing interests:** The authors declare no competing interests.

## Abstract

**Background:** While general population studies have shown associations between greater physical activity and lower inflammatory markers, effects of physical activity on inflammatory pathways in rheumatoid arthritis (RA) remain unknown. We aimed to determine whether physical activity independently associates with differential expression of inflammatory genes in RA.

**Methods:** Data derived from an observational RA cohort. Physical activity was measured with the GT9X ActiGraph Link device. RNA extraction from peripheral blood, sequencing library preparation and transcriptomic analyses were performed using established methods. Genes differentially expressed in the most versus least physically active groups (top versus bottom activity tertiles) were identified using DESeq2 after adjusting for sex, age, race/ethnicity, and disease activity. Ingenuity Pathway Analysis (IPA) was employed to identify canonical biological pathways and upstream regulating cytokines associated with physical activity.

**Results:** 35 participants were enrolled (mean age 56±12 years; 91% female; race/ethnicity 31% white, 9% African American, 9% Asian, 40% Hispanic). 767 genes were differentially expressed (padj<0.1) between high versus low activity groups. The high activity group exhibited downregulation of innate and adaptive immune signaling pathways, including CD40, STAT3, TREM-1, IL-17a, IL-8, toll-like receptor and interferon signaling. Upstream cytokine analysis demonstrated inhibition of TNF-alpha and interferon among individuals in the most active group.

**Conclusion:** In a racially diverse RA cohort, patients who were more physically active had lower expression of immune signaling pathways implicated in RA pathogenesis, even after adjusting for disease activity, suggesting a potential protective effect of physical activity in RA.

## Introduction

Rheumatoid arthritis (RA) is a systemic autoimmune disease that affects 1.5 million Americans and is characterized by joint and systemic inflammation, resulting in chronic pain, functional limitations, and premature cardiovascular disease.^1^ While existing therapeutics such as biologic disease-modifying antirheumatic drugs (DMARDs) reduce disease severity, they often fail to adequately control symptoms and can lead to life-threatening adverse events. Given these limitations, there is an unmet need for non-pharmacologic strategies to augment existing medical management of RA, improve long-term clinical outcomes, and reduce disease-related symptoms.

While historically it was thought that exercise might exacerbate rheumatic conditions, that precept has since been disproven.^2^ Driving this shift were clinical trials of resistance and aerobic exercise programs for people with RA, in which the exercise treatment interventions were found to be safe and resulted in unchanged or improved disease activity scores compared to controls.^3,4^ As an example, a recent exercise pilot trial found that older adults with RA randomized to a 10-week high-intensity interval walking intervention had a 38% reduction in disease activity at the end of the intervention.^5^

The mechanisms responsible for the therapeutic effect of exercise in rheumatic diseases are incompletely understood. One hypothesis is that skeletal muscle-secreted myokines confer anti-inflammatory effects following episodes of physical activity.^6^ This hypothesis is supported by a number of studies in the general population that demonstrated a clear association between higher levels of physical activity, reductions in pro-inflammatory cytokine signaling, and lower systemic inflammation.^7–10^ Though studies in the general population have observed favorable effects of exercise on immune function, the impact of physical activity on systemic inflammation in the context of immune-mediated inflammatory diseases such as RA remains poorly understood and in need of further investigation.

Here we conduct a prospective observational cohort study to investigate mechanistic relationships between physical activity and systemic inflammatory gene expression in patients with RA. Using quantitative actigraphy, whole blood transcriptomics, and detailed clinical phenotyping, we ask whether physical activity independently associates with expression of inflammatory genes implicated in the pathogenesis of RA.

## Methods

### Study design, clinical cohort and ethics statement

We studied participants enrolled in an ongoing prospective observational cohort study of patients with RA: “Sleep disturbance in rheumatoid arthritis: phenotypes, causes, and impact” (RAZZ; R01 AR069616). Patients were enrolled sequentially between 2018 and 2020 after being recruited from an extensive network of community, academic, and safety net rheumatology clinics in the San Francisco Bay Area, including the University of California San Francisco (UCSF) and Zuckerberg San Francisco General (ZSFG) Hospital rheumatology clinics. Inclusion criteria for this study were: age greater than or equal to 18 years; a physician’s diagnosis of RA; and English or Spanish fluency.

During the period of data collection for this analysis, study visits were conducted in-person. At each study visit, we collected detailed clinical information, assessed RA disease activity, and downloaded actigraphy data collected between study visits. In addition, peripheral blood was collected and stored at −80°C for subsequent RNA sequencing (RNA-seq). The UCSF Institutional Review Board approved all procedures (UCSF IRB protocol #17-21790) and all participants provided written informed consent.

### Clinical measures

#### Physical activity

Physical activity was assessed objectively using the GT9X ActiGraph Link device (ActiGraph, Inc). This ActiGraph device resembles a small watch and contains a validated tri-axial accelerometer and integrated gyroscope and magnetometer, which collectively capture absolute subject movement. Participants wore the ActiGraph on their wrists for 7 consecutive 24-hour periods (one full week) before returning them for data analysis. ActiGraph physical activity data are provided as counts per minute (CPM), which are a result of aggregating post-filtered raw accelerometer data over 1-minute intervals (epochs) and are used to define the time awake spent in sedentary activity (0-99 CPM), light activity (100-1951 CPM), moderate activity (1952-5724 CPM), vigorous activity (5725-9498 CPM), and very vigorous activity (>=9499 CPM) based on established cut points that correspond to metabolic equivalent (MET) levels.^11^ Participants were categorized using the highest, middle, and lowest tertiles for percentage of time spent in at least moderately intense physical activity across the entire RA cohort, resulting in three physical activity groups: the least active (labeled “inactive”), the most active (labeled “active”), and an intermediate group.

#### RA-specific disease factors

Age of diagnosis was obtained by self-report. Disease activity was assessed with the Rheumatoid Arthritis Disease Activity Index (RADAI), a validated patient-reported instrument.^12^ Participants were also queried regarding current treatment with glucocorticoids—including dosage and frequency—as well as other immunomodulatory medications.

#### Other variables

Sociodemographic data was collected, including sex, age, race, ethnicity, and educational attainment (categorized as high school graduate or less, versus those with additional education). Height and weight were measured during the baseline in-person visit, and body mass index (BMI) was calculated as weight in kilograms (kg) divided by height in meters squared (m^2^). Participants provided information on health-related behaviors, such as smoking, and comorbidities such as cardiovascular disease, diabetes mellitus, asthma, and cancer.

### RNA sequencing

Whole blood was collected and stored at −80C. Following RNA extraction (Zymo Pathogen Magbead Kit) and DNAse treatment, human globin and ribosomal RNA was depleted using FastSelect (Qiagen) according to described methods.^13^ RNA was then fragmented and underwent library preparation using the NEBNext Ultra II RNAseq Kit (New England Biolabs) as previously described.^13^ Libraries underwent 146 nucleotide paired-end Illumina sequencing on an Illumina Novaseq 6000 instrument.

### Gene expression data processing and quality control

Following demultiplexing, sequencing reads were aligned against the human genome (NCBI GRC h38) using STAR^4^ to extract gene counts. Samples retained in the dataset had ≥ 3.0 × 10^5^ gene counts, and the median across all samples was 5.8 × 10^5^ gene counts. Data merging and normalization across different activity groups were performed. Gene counts were normalized with the median of ratios method using R package *DEseq2*. For covariates, one-hot encoding was applied for categorical variables and min-max scaling was performed for numeric covariates. As an additional quality control measure, genes expressed in less than 30 percent of the patients in each group were filtered. In total 19,260 genes were kept for the subsequent analysis.

### Differential gene expression analysis

Differentially expressed genes were identified using the R package *DEseq2*.^14^ Sex, age, race, and ethnicity were included as covariates in the linear model. To correct for the potential batch effect, the R package *sva*^15^ was used to calculate a surrogate variable (SV), which was also integrated into the differential expression linear model as a covariate. Independent hypothesis weighting (IHW) was used as a multiple testing procedure and the significance of differential expression was defined as an adjusted p-value < 0.1.

### Pathway analysis

Ingenuity Pathway Analysis (IPA)^16^ as carried out on differentially expressed genes with a P < 0.1 ranked by log2 fold change. Significant IPA results were defined as those with a Z-score absolute value greater than 2 or an overlap P value < 0.05. The top three up- and down-regulated canonical pathways based on Z-score, as well as all pathways related to immunity and inflammation with an |Z| > 1 and overlap P value < 0.05, were included in Figure 1C. Upstream regulating cytokines with an |Z| > 2 and overlap P value < 0.05 were included in Figure 1D. Complete IPA results are provided in supplementary materials (**Supplementary Data File 2**).

**Figure 1.**
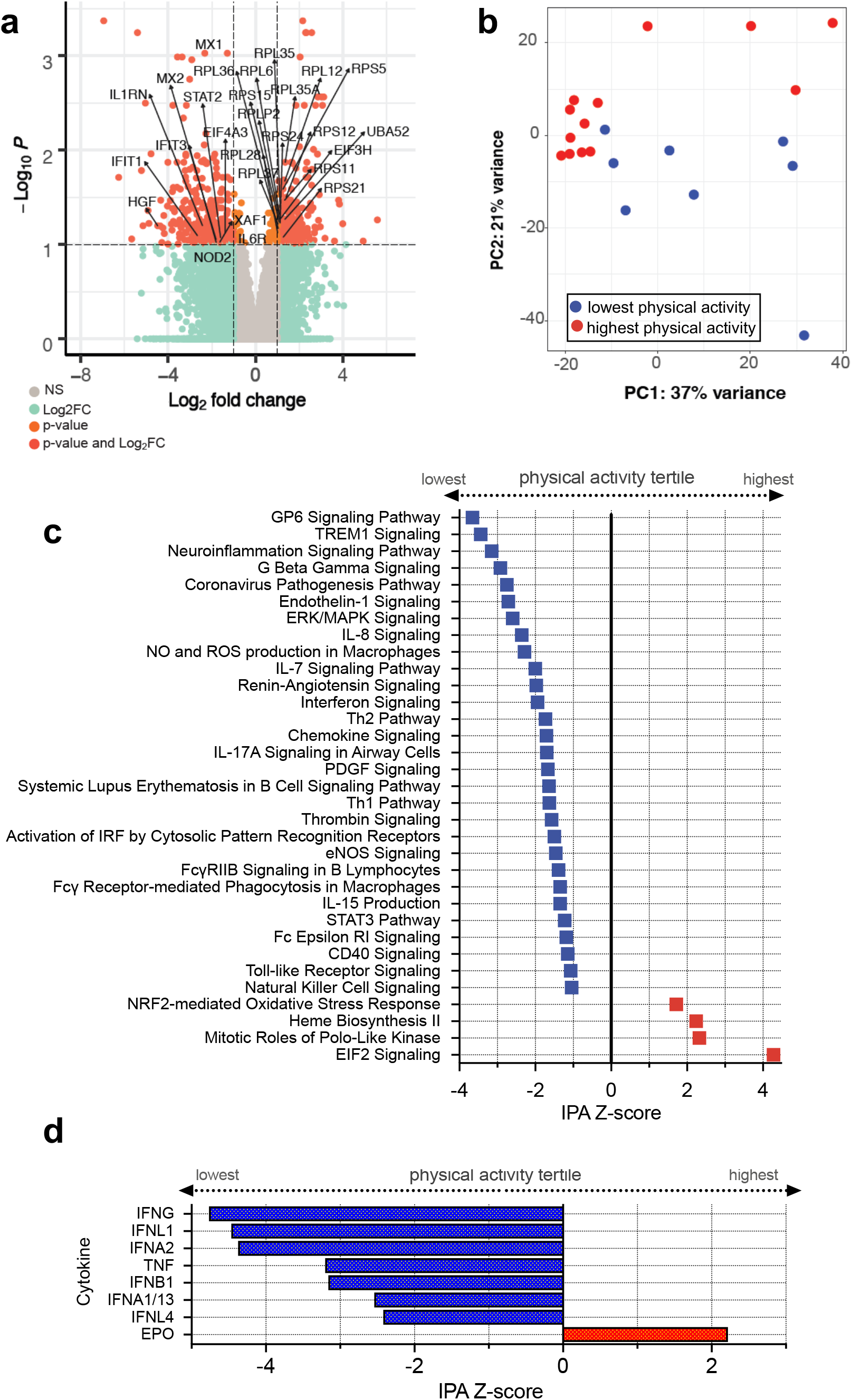
Differentially expressed (DE) genes in peripheral blood between RA patients in the highest and lowest tertiles of physical activity. **a)** Volcano plot of DE genes depicting 365 genes up-regulated in the highest activity tertile and 402 down-regulated genes at an adjusted p-value < 0.1 with respect to the highest physical activity tertile. Genes related to immune signaling and translation are highlighted. **b)** Principal Component Analysis based on the DE genes demonstrates separation of patients based on tertile of physical activity. **c)** Ingenuity Pathway Analysis (IPA) based on differential gene expression analyses demonstrating expression of canonical signaling pathways by IPA activation Z-score. Top pathways by |Z| score and pathways related to immunity and inflammation with a |Z| > 1 depicted, with an overlap P value < 0.05. Values and related genes tabulated in (Supplementary Data X). **d)** Predicted activation state of upstream cytokines in the highest versus lowest physical activity tertiles. Cytokines with a |Z| > 2 plotted. Values and related genes tabulated in (Supplementary Data X). **e)** *In silico* cell type enrichment analysis depicting differences in peripheral immune cell populations based on physical activity tertile.

### Network analysis

The network analysis was performed using STRING v.11. (https://string-db.org/) The network matrix was exported as a .tsv file and the plot was recreated using Cytoscape (https://cytoscape.org/). Each protein was visualized as a node and each potential protein-protein interaction was visualized as an edge. The size of each node reflects its degree centrality metrics. **Figure 2A** represents one of multiple network maps. Complete network map data are provided in **Supplementary Data File 3**.

**Figure 2.**
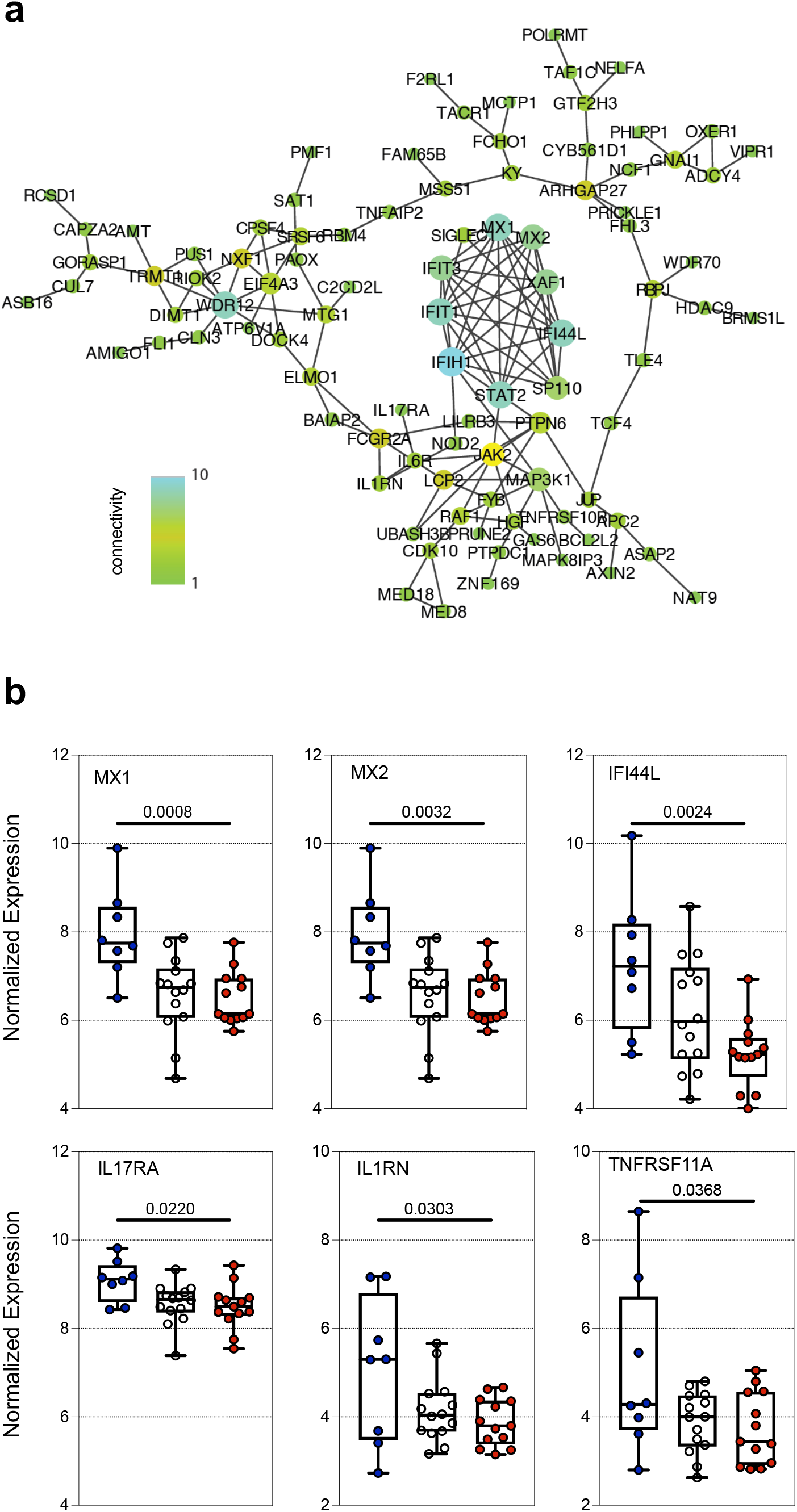
Connectivity and dose-dependent downregulation of proinflammatory genes based on physical activity tertile. a) Network connectivity map of proinflammatory genes downregulated in the highest physical activity tertile, constructed using STRING v.11. Color bar represents the degree of connectivity based on the number of edges to which each node/gene is connected. Complete network connectivity table of all differentially expressed genes in **Supplementary Data File 3**. b) Boxplots depicting normalized expression of six representative immune genes correlated with physical activity tertile in a dose-dependent manner.

### *In silico* analysis of cell type proportions

Cell-type proportions were estimated from bulk host transcriptome data using the CIBERSORT X algorithm following previously described methods.^13^ The estimated proportions were compared between the three patient groups using a Mann-Whitney-Wilcoxon test (two-sided) with Bonferroni correction.

### Data Availability

Genecounts are available under Gene Expression Omnibus accession number GSE179302. All code is available at Github https://github.com/drychkov/PA_in_RA.

## Results

There were 35 adult RA patients in the cohort with actigraphy and metagenomic data available for analysis. The cohort had a mean age of 56 years (SD 12.1), was 91% female, and self-reported the following racial and ethnic identities: 31% white, 9% African American, 9% Asian, and 40% Hispanic (**Table 1**). The mean disease duration was 13.1 years (SD 11.3), 54% were taking methotrexate, 51% were treated with a biologic DMARD, and 26% were treated with systemic glucocorticoids. Only 13% were taking oral glucocorticoids greater than 7.5 mg prednisone equivalent per day. None of the patients engaged in vigorous physical activity during the 7-day period of actigraphy monitoring. The percentage of time spent in moderate physical activity was 18% in the most active group compared to 4.5% in the least active group (**Supplementary Table 1**). Patients in the active group were younger (mean age = 50 years) compared to those in the intermediate (mean age = 56 years) and inactive groups (mean age = 63 years) (p = 0.04), but there were no other statistically significant demographic or clinical differences between physical activity groups (**Table 1**).

**Table 1.** Characteristics* of Patients with Rheumatoid Arthritis (RA) According to Physical Activity Category.

| Characteristics | Overall<br>(N = 35) | Physical Activity** Status |  |  | P |
| --- | --- | --- | --- | --- | --- |
|  |  | Active<br>(N = 13) | Intermediate<br>(N=14) | Inactive<br>(N=8) |  |
| <u>Sociodemographic Factors:</u> |  |  |  |  |  |
| Age, mean ± SD | 56 ± 12 | 50 ± 11 | 56 ± 11 | 63 ± 12 | **0.04** |
| Female | 32 (91%) | 13 (100%) | 13 (93%) | 6 (75%) | 0.06 |
| Race |  |  |  |  | 0.21 |
| Asian | 3 (9%) | 0 (0%) | 2 (14%) | 1 (13%) |  |
| African American | 3 (9%) | 0 (0%) | 1 (7%) | 2 (25%) |  |
| White | 11 (31%) | 3 (23%) | 4 (29%) | 4 (50%) |  |
| Unknown | 14 (40%) | 8 (62%) | 5 (36%) | 1 (13%) |  |
| Multiple | 4 (11%) | 2 (15%) | 2 (14%) | 0 (0%) |  |
| Hispanic ethnicity | 14 (40%) | 8 (62%) | 5 (36%) | 1 (13%) | 0.08 |
| Education < college degree | 18 (51%) | 8 (62%) | 6 (43%) | 5 (50%) | 0.62 |
| <u>RA Specific Characteristics:</u> |  |  |  |  |  |
| RA disease duration, mean years ± SD | 13 ± 11 | 12 ± 11 | 14 ± 13 | 15 ± 12 | 0.82 |
| Disease activity by RADAI, mean ± SD | 3.9 + 1.9 | 3.9 + 1.3 | 4.5 + 2.1 | 3.1 + 1.8 | 0.21 |
| Treated with methotrexate | 19 (54%) | 8 (62%) | 5 (36%) | 6 (75%) | 0.17 |
| Treated with TNFi | 12 (34%) | 3 (23%) | 7 (50%) | 2 (25%) | 0.28 |
| Treated with any conventional DMARD | 24 (69%) | 9 (69%) | 8 (57%) | 7 (88%) | 0.34 |
| Treated with any biologic DMARD | 18 (51%) | 6 (46%) | 10 (71%) | 2 (25%) | 0.10 |
| Current systemic glucocorticoid use | 9 (26%) | 3 (23%) | 4 (29%) | 2 (25%) | 0.95 |
| <u>Comorbidities and Health Status:</u> |  |  |  |  |  |
| Cardiovascular Disease | 2 (6%) | 0 (0%) | 1 (7%) | 1 (13%) | 0.47 |
| Diabetes Mellitus | 5 (14%) | 1 (8%) | 3 (21%) | 1 (13%) | 0.59 |
| Asthma | 5 (14%) | 2 (15%) | 1 (7%) | 2 (25%) | 0.51 |
| History of malignancy | 3 (9%) | 0 (0%) | 1 (7%) | 2 (25%) | 0.14 |
| Body Mass Index (kg/m2), mean ± SD | 28.8 ± 5.5 | 29.0 ± 6.8 | 28.9 ± 5.2 | 28.6 ± 4.1 | 0.33 |
| Current nicotine use | 2 (6%) | 1 (8%) | 0 (0%) | 1 (13%) | 0.44 |

\*Data are n (%) unless otherwise indicated. P-values calculated using chi-squared tests for categorical measures and ANOVA for continuous measures.
\*\*Physical activity groups were created from the lower (inactive), middle (intermediate), and upper (active) tertiles for percentage of time spent in at least moderately intense physical activity among all study participants in the larger RA cohort.
RADAI = Rheumatoid Arthritis Disease Activity Index, range 0-10.
TNFi = tumor necrosis factor inhibitor.
DMARD = disease modifying antirheumatic drug.
Cadiovascular disease was defined by history of stroke, coronary artery disease, and/or myocardial infarction.

Differential gene expression analysis comparing the most versus least physically active groups (based on activity tertile) identified 767 genes at an adjusted P value (padj) < 0.1 (**Figure 1a**). Principal Component Analysis (PCA) demonstrated clear separation based on physical activity groups (**Figure 1b**), and pathway analysis (**Methods**) revealed that the most physically active patients exhibited downregulation of diverse innate and adaptive immune signaling pathways implicated in the pathogenesis of RA, including CD40, STAT3, TREM-1, IL-17a, IL-8, toll-like receptors, and type I interferon signaling (**Figure 1c, Supplementary Data File 1, 3A)**.^17^

Prediction of upstream cytokine activation states from the transcriptomic data suggested inhibition of type I, II and III interferons, and activation of epoetin, among the most physically active patients (**Figure 1d, Supplementary Data File 2B**).

Given the role of pathologic inflammation in RA, we next sought to more rigorously evaluate the proinflammatory genes downregulated in the most active group by performing a network connectivity analysis. This revealed relationships between genes related to type 1 interferon signaling (e.g. *MX1, IFI44L, IFIT1*) and inflammasome signaling (e.g. *IL-1RN, NOD2)*, as well as other cytokine signaling pathways (e.g. *IL-6R, IL17RA)* (**Figure 2a, Supplementary Data File 3**). To more deeply characterize the relationships between these inflammatory genes and physical activity, we evaluated their expression across all three physical activity groups. Intriguingly, a dose-dependent correlation between physical activity and reduced expression of proinflammatory genes was observed (**Figure 2b**).

We considered that differences in immune cell populations might underlie the observed differentially expressed genes, and thus performed *in silico* cell type deconvolution. We found no statistically significant differences in predicted proportions between the highest and lowest physical activity groups, but we observed a trend toward decreased monocytes in the most active group (**Supplementary Figure 1, Supplementary Data File 4**). Lastly, we considered that differences in ability to engage in exercise among patients with more versus less severe/active disease might explain our findings. To test for this possibility, we conducted a sensitivity analysis in which disease activity was included as a covariate. The sensitivity analysis revealed consistent results compared to the main analysis, including 674 differentially expressed genes (padj < 0.1) and similar downregulation of interferon and other proinflammatory cytokine signaling pathways among the most active patients (**Supplementary Data File 4**).

## Discussion

Randomized controlled trials of exercise interventions for RA have found that exercise improves joint pain, fatigue, and disease activity,^3,4,18–21^ and decreases the risk of progressive joint damage^18^, but the mechanisms underlying these apparent benefits have remained in question. Here we find that physical activity moderates inflammatory signaling in RA patients using quantitative actigraphy and whole blood transcriptomics.

Studies in the general population have found that higher levels of physical activity associate with lower inflammatory biomarkers, including C-reactive protein (CRP) and TNF-a, even after adjusting for excess adiposity ^7–10^. Our results are consistent with these observations, as we observed an association between physical activity and attenuation of several proinflammatory signaling pathways implicated in RA pathogenesis, including CD40, STAT3, TREM-1, IL-17a, IL-8, toll-like receptor, and type I interferon signaling.^17^ Furthermore, our gene expression data demonstrated lower TNF activation among the most active patients in this RA cohort.

Unexpectedly, we found that interferon signaling was inversely correlated with physical activity. This finding has potential clinical significance given that interferon gene expression in peripheral blood has been associated with RA autoantibody production, development of chronic RA in patients with early inflammatory arthritis, poorer response to initial therapy, and nonresponse to rituximab^22^. Additionally, recent single cell RNA-seq studies have shown a correlation between peripheral blood mononuclear cell interferon gene expression and plasma cell infiltration of synovium in people with RA^23^. We also found that physical activity was associated with significantly attenuated toll-like receptor and *IL-17RA* signaling, additional pathways implicated in RA pathogenesis.^24^ We considered that disease severity might represent a potential confounder, but in a sensitivity analysis that included disease activity as a covariate, we found consistent results.

Intriguingly, physical activity was associated with reduced expression of several proinflammatory genes in a dose-dependent manner. The threshold of moderate activity employed in Actigraph scoring is quite modest at ∼2.0 METs, which corresponds to home activities such as standing to wash dishes, food shopping, or walking at a ≤2.0 mph pace. The most active group spent an average of 4.3 hours/day in these moderate intensity activities, compared to the least active group’s 1.1 hours/day. The dose-dependent relationship between moderate activity and attenuation of proinflammatory gene expression suggests that greater exposure to physical activity confers a larger impact on inflammatory signaling—and therefore greater potential therapeutic benefit—in this patient population.

This study has limitations. Because this was an observational study, we can only identify an association between gene expression and physical activity, but not prove causation or directionality. We also examined only a limited group of patients in this first study to assess physical activity in the context of RA using whole blood transcriptomics. A future randomized clinical trial will be needed to assess the direct effect of exercise on blood transcriptomic markers. We were unable to assess the impact of vigorous physical activity because no patient engaged in vigorous physical activity during the study, which may have attenuated the underlying signal. However, this may also be considered an advantage, as our data realistically reflect physical activity patterns observed in this patient demographic.^25^ If anything, our results may have been more pronounced in a cohort with a broader range of activity levels.

Given the limitations of existing pharmacologic treatments for RA and the need for non-pharmacologic adjunctive strategies, our findings have important clinical implications. Despite major advances in treatment, including the advent of biologic disease modifying antirheumatic drugs (DMARDs), RA remains a chronic, incurable condition, and fewer than half of patients treated with immunosuppressive DMARDs achieve disease remission.^26,27^ Furthermore, adverse drug events from RA medications are common, with an estimated incidence of 15 per 100 patient-years,^28^ leading many people with RA to express a preference for non-pharmacologic treatment approaches.^29,30^ Taken together, the prevalence of persistent symptoms despite treatment, risk of side effects from DMARDs, and patient preference for lower-risk treatment approaches emphasize the critical need for adjunctive non-pharmacologic interventions for RA. Our results suggest that physical activity interventions have the potential to not only improve overall health and mitigate the risk of important comorbidities among people with RA, but may also attenuate pathologic inflammatory signaling and disease activity.

This work provides an important foundation for future studies. There is an important opportunity for additional research to further characterize the biological mechanisms linking physical activity to inflammatory signaling using proteomic, metabolomic, and single cell RNA sequencing approaches. It will be important to validate our findings in the context of a randomized controlled trial (RCT) of exercise for people with RA, and to compare gene expression among RA patients across a greater spectrum of physical activity. Lastly, a key outstanding question is whether our findings extend to other autoimmune rheumatic diseases such as systemic lupus erythematosus (SLE).

In summary, among a representative cohort of RA patients, we found a striking and dose-dependent association between moderate physical activity and attenuated expression of genes involved in both innate and adaptive immune signaling, even after adjusting for disease severity and other covariates. These findings provide the first mechanistic evidence to support a disease-modifying effect of physical activity in RA.

## Acknowledgements

This work is supported by NIH/NIAMS R01 AR069616 and NIH/NHLBI K23HL138461-01A1. Additional support from the Rheumatology Research Foundation.

## References

1 Hunter, T. M. et al. Prevalence of rheumatoid arthritis in the United States adult population in healthcare claims databases, 2004-2014. Rheumatol. Int. 37, 1551–1557, doi:10.1007/s00296-017-3726-1 (2017).

2 Lundberg, I. E. & Nader, G. A. Molecular effects of exercise in patients with inflammatory rheumatic disease. Nat. Clin. Pract. Rheumatol. 4, 597–604, doi:10.1038/ncprheum0929 (2008).

3 Baillet, A. et al. Efficacy of cardiorespiratory aerobic exercise in rheumatoid arthritis: meta-analysis of randomized controlled trials. Arthritis Care Res. (Hoboken) 62, 984–992, doi:10.1002/acr.20146 (2010).

4 Conn, V. S., Hafdahl, A. R., Minor, M. A. & Nielsen, P. J. Physical activity interventions among adults with arthritis: meta-analysis of outcomes. Semin. Arthritis Rheum. 37, 307–316, doi:10.1016/j.semarthrit.2007.07.006 (2008).

5 Bartlett, D. B. et al. Ten weeks of high-intensity interval walk training is associated with reduced disease activity and improved innate immune function in older adults with rheumatoid arthritis: a pilot study. Arthritis Res. Ther. 20, 127, doi:10.1186/s13075-018-1624-x (2018).

6 Benatti, F. B. & Pedersen, B. K. Exercise as an anti-inflammatory therapy for rheumatic diseases-myokine regulation. Nat. Rev. Rheumatol. 11, 86–97, doi:10.1038/nrrheum.2014.193 (2015).

7 Abramson, J. L. & Vaccarino, V. Relationship between physical activity and inflammation among apparently healthy middle-aged and older US adults. Arch. Intern. Med. 162, 1286–1292 (2002).

8 Ford, E. S. Does exercise reduce inflammation? Physical activity and C-reactive protein among U.S. adults. Epidemiology 13, 561–568, doi:10.1097/01.EDE.0000023965.92535.C0 (2002).

9 Colbert, L. H. et al. Physical activity, exercise, and inflammatory markers in older adults: findings from the Health, Aging and Body Composition Study. J. Am. Geriatr. Soc. 52, 1098–1104, doi:10.1111/j.1532-5415.2004.52307.x (2004).

10 Lavie, C. J., Church, T. S., Milani, R. V. & Earnest, C. P. Impact of physical activity, cardiorespiratory fitness, and exercise training on markers of inflammation. J. Cardiopulm. Rehabil. Prev. 31, 137–145, doi:10.1097/HCR.0b013e3182122827 (2011).

11 Freedson, P. S., Melanson, E. & Sirard, J. Calibration of the Computer Science and Applications, Inc. accelerometer. Med. Sci. Sports Exerc. 30, 777–781, doi:10.1097/00005768-199805000-00021 (1998).

12 Hendrikx, J., de Jonge, M. J., Fransen, J., Kievit, W. & van Riel, P. L. Systematic review of patient-reported outcome measures (PROMs) for assessing disease activity in rheumatoid arthritis. RMD Open 2, e000202, doi:10.1136/rmdopen-2015-000202 (2016).

13 Mick, E. et al. Upper airway gene expression reveals suppressed immune responses to SARS-CoV-2 compared with other respiratory viruses. Nat Commun 11, 5854, doi:10.1038/s41467-020-19587-y (2020).

14 Love, M. I., Huber, W. & Anders, S. Moderated estimation of fold change and dispersion for RNA-seq data with DESeq2. Genome Biol. 15, 550, doi:10.1186/s13059-014-0550-8 (2014).

15 Leek, J. T., Johnson, W. E., Parker, H. S., Jaffe, A. E. & Storey, J. D. The sva package for removing batch effects and other unwanted variation in high-throughput experiments. Bioinformatics 28, 882–883, doi:10.1093/bioinformatics/bts034 (2012).

16 Kramer, A., Green, J., Pollard, J., Jr. & Tugendreich, S. Causal analysis approaches in Ingenuity Pathway Analysis. Bioinformatics 30, 523–530, doi:10.1093/bioinformatics/btt703 (2014).

17 Noack, M. & Miossec, P. Selected cytokine pathways in rheumatoid arthritis. Semin. Immunopathol. 39, 365–383, doi:10.1007/s00281-017-0619-z (2017).

18 de Jong, Z. et al. Long term high intensity exercise and damage of small joints in rheumatoid arthritis. Ann. Rheum. Dis. 63, 1399–1405, doi:10.1136/ard.2003.015826 (2004).

19 Lyngberg, K., Danneskiold-Samsoe, B. & Halskov, O. The effect of physical training on patients with rheumatoid arthritis: changes in disease activity, muscle strength and aerobic capacity. A clinically controlled minimized cross-over study. Clin. Exp. Rheumatol. 6, 253–260 (1988).

20 Neuberger, G. B. et al. Effects of exercise on fatigue, aerobic fitness, and disease activity measures in persons with rheumatoid arthritis. Res. Nurs. Health 20, 195–204, doi:10.1002/(sici)1098-240x(199706)20:3<195::aid-nur3>3.0.co;2-d (1997).

21 van den Ende, C. H. et al. Effect of intensive exercise on patients with active rheumatoid arthritis: a randomised clinical trial. Ann. Rheum. Dis. 59, 615–621, doi:10.1136/ard.59.8.615 (2000).

22 Rodriguez-Carrio, J., Lopez, P. & Suarez, A. Type I IFNs as biomarkers in rheumatoid arthritis: towards disease profiling and personalized medicine. Clin. Sci. (Lond.) 128, 449–464, doi:10.1042/CS20140554 (2015).

23 Lewis, M. J. et al. Molecular Portraits of Early Rheumatoid Arthritis Identify Clinical and Treatment Response Phenotypes. Cell Rep 28, 2455–2470 e2455, doi:10.1016/j.celrep.2019.07.091 (2019).

24 Kehlen, A., Thiele, K., Riemann, D. & Langner, J. Expression, modulation and signalling of IL-17 receptor in fibroblast-like synoviocytes of patients with rheumatoid arthritis. Clin. Exp. Immunol. 127, 539–546, doi:10.1046/j.1365-2249.2002.01782.x (2002).

25 Sokka, T. et al. Physical inactivity in patients with rheumatoid arthritis: data from twenty-one countries in a cross-sectional, international study. Arthritis Rheum. 59, 42–50, doi:10.1002/art.23255 (2008).

26 Hetland, M. L. et al. Direct comparison of treatment responses, remission rates, and drug adherence in patients with rheumatoid arthritis treated with adalimumab, etanercept, or infliximab: results from eight years of surveillance of clinical practice in the nationwide Danish DANBIO registry. Arthritis Rheum. 62, 22–32, doi:10.1002/art.27227 (2010).

27 van der Heijde, D. et al. Disease remission and sustained halting of radiographic progression with combination etanercept and methotrexate in patients with rheumatoid arthritis. Arthritis Rheum. 56, 3928–3939, doi:10.1002/art.23141 (2007).

28 Abasolo, L. et al. Safety of disease-modifying antirheumatic drugs and biologic agents for rheumatoid arthritis patients in real-life conditions. Semin. Arthritis Rheum. 44, 506–513, doi:10.1016/j.semarthrit.2014.11.003 (2015).

29 Neame, R. & Hammond, A. Beliefs about medications: a questionnaire survey of people with rheumatoid arthritis. Rheumatology (Oxford) 44, 762–767 (2005).

30 Palominos, P. E. et al. Fears and beliefs of people living with rheumatoid arthritis: a systematic literature review. Adv Rheumatol 58, 1 (2018).

