## Supplementary Materials for "Physical Activity Moderates Inflammatory Gene Expression in Rheumatoid Arthritis"

**Supplementary Material**

**Supplementary Table 1. Physical Activity[1] Behavior among RA Patients by Physical Activity Groups**

| | Overall | Physical Activity[2] Groups | | |
|---|---|---|---|---|
| | | Inactive | Intermediate | Active |
| | (N = 35) | (N = 8) | (N=14) | (N = 13) |
| **Absolute time in each type of activity** | | | | |
| Sedentary activity, mean hours/day ± SD | 11.4 ± 2.0 | 13.4 ± 1.9 | 11.3 ± 1.8 | 10.4 ± 1.3 |
| Light activity, mean hours/day ± SD | 9.8 ± 1.7 | 9.6 ± 2.0 | 10.4 ± 1.7 | 9.3 ± 1.4 |
| Moderate activity, mean hours/day ± SD | 2.8 ± 1.5 | 1.1 ± 0.4 | 2.3 ± 0.3 | 4.3 ± 1.1 |
| Vigorous activity, mean hours/day ± SD | 0.0 | 0.0 | 0.0 | 0.0 |
| **Percent time in each type of activity** | | | | |
| Sedentary activity[3], % | 47.6% | 55.6% | 47.2% | 43.2% |
| Light activity[4], % | 40.8% | 39.9% | 43.3% | 38.8% |
| Moderate activity[5], % | 11.6% | 4.5% | 9.6% | 18.0% |
| Vigorous activity[6], % | 0.0% | 0.0% | 0.0% | 0.0% |

[1]Physical activity measured with the GT9X ActiGraph Link device, worn for 7 consecutive 24-hour periods.

[2]Physical activity groups were categorized by lower (inactive), middle (intermediate), and upper (active) tertiles for percentage of time spent in at least moderately intense physical activity among all study participants.

[3]Sedentary activity: 0-99 counts per minute, metabolic equivalent (MET) level ≤1.5

[4]Light activity: 100-1951 counts per minute, MET level 1.51-2.99

[5]Moderate activity: 1952-5724 counts per minute, MET level 3.0-5.99 METS

[6]Vigorous activity: ≥ 5725 counts per minute, MET level ≥6.0 METS

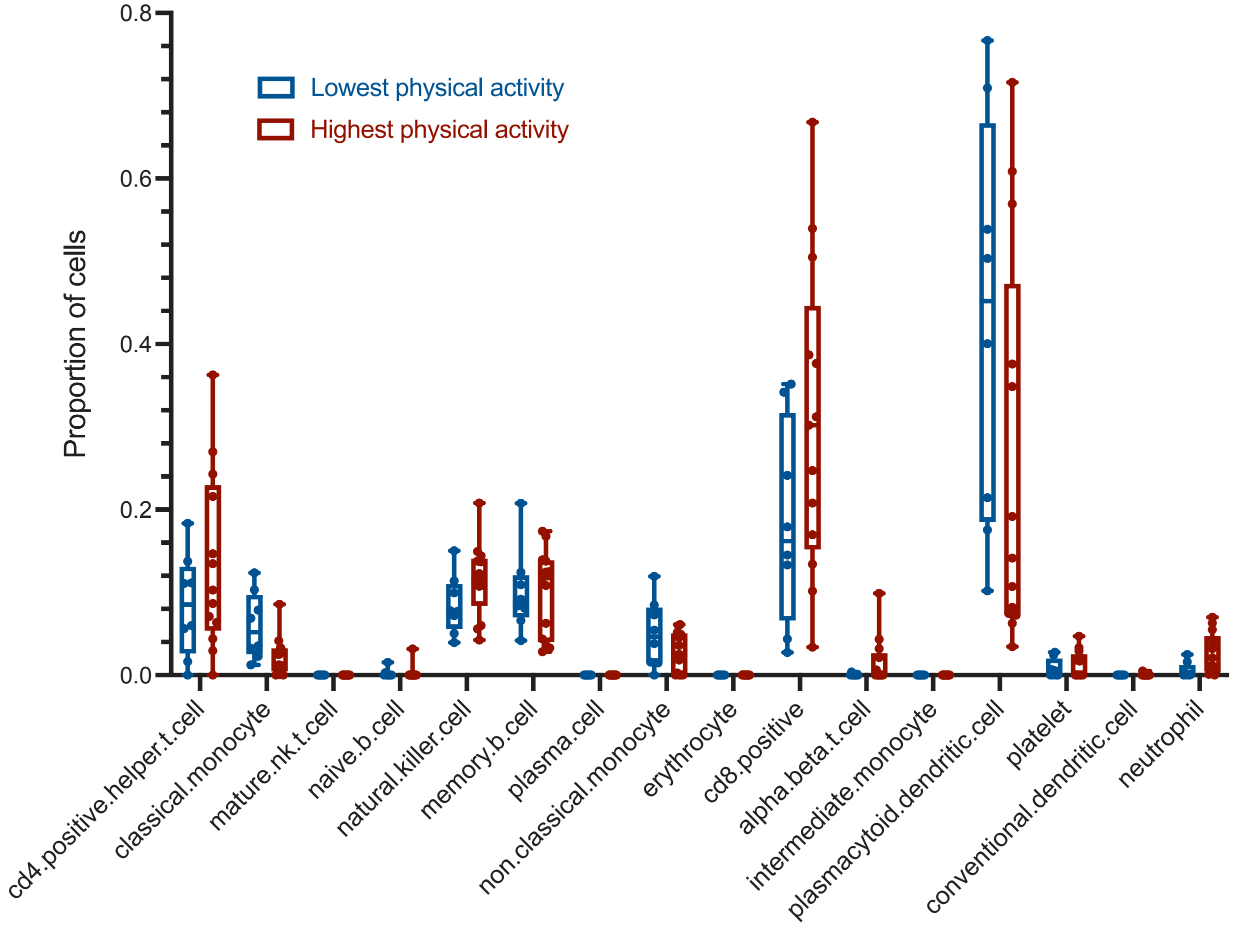


**Supplementary Figure 1. Cell type proportion differences between RA patients with the highest and lowest tertiles of physical activity.** *In silico* deconvolution of cell type proportions from whole blood RNA-sequencing Data File using blood single cell signatures. Pairwise comparisons between patient groups were performed with a two-sided Mann-Whitney-Wilcoxon test followed by Bonferroni's correction. No statistically significant differences in cell type proportions were identified.

**Supplementary Data Files**

Supplementary Data File 1. Differentially expressed genes (padj < 0.1) between the highest and lowest tertiles of physical activity. Sex, age, race, and ethnicity were included as covariates in the linear model.

Supplementary Data File 2. A. Ingenuity Pathway Analysis (IPA) canonical pathways. B. IPA predicted activation states of upstream regulating cytokines.

Supplementary Data File 3. Complete network connectivity analysis of differentially expressed genes between the highest and lowest tertiles of physical activity. A. Upregulated gene networks. B. Downregulated gene networks.

Supplementary Data File 4. *In silico* prediction of cell type proportions from transcriptomic data.

Supplementary Data File 5. Differentially expressed genes after also including RADAI as a covariate, in addition to sex, age, race, and ethnicity.
